# Wavelet analysis of climate variability and malaria incidence to inform intervention planning in low- and high-burden Nigerian states

**DOI:** 10.64898/2026.02.28.26347314

**Authors:** Samuel Abidemi Osikoya, Emmanuel Afolabi Bakare, Lukman Shina Akinola, Olusola Oresanya, Chukwu Okoronkwo, Eze Nelson, Ibrahim Maikore

**Author notes:** These authors contributed equally to this work (Samuel Abidemi Osikoya, Emmanuel Afolabi Bakare). These authors also contributed equally to this work (Lukman Shina Akinola, Olusola Oresanya, Chukwu Okoronkwo, Eze Nelson, Ibrahim Maikore).

## Abstract

Malaria remains a major public health challenge in Nigeria, and increasing climate variability poses substantial threats to recent gains in control. However, malaria transmission does not respond uniformly to climate drivers across epidemiological settings, highlighting the need to explore climate-malaria dynamics within heterogeneous contexts. This study examined the non-stationary temporal dynamics of malaria incidence and two key climatic drivers—rainfall and temperature—in Lagos and Zamfara states. These states were selected to represent heterogeneous transmission intensities, urbanisation and climatic regimes. Monthly malaria incidence and corresponding climate data (2015–2024) were analysed using wavelet-based model to characterise the non-stationary periodicities, quantify time-varying climate–malaria associations and identify time-dependent lead–lag relationships. Malaria incidence exhibited transient semi-annual, annual, and multi-annual cycles that were weak and temporally localized, despite persistent annual cycles in rainfall and temperature in Lagos. Cross-wavelet spectra revealed intermittent associations within the 8–16-month band, while phase analysis indicated short-lived alignment in which malaria incidence lagged rainfall by approximately one month, particularly between 2019 and 2022. The relationship with temperature was unstable, suggesting rainfall exerted more consistent influence on malaria incidence. In contrast, Zamfara displayed strong and dominant annual cycles of malaria incidence throughout the study period, with rainfall and temperature showing stable, statistically significant annual co-variability. Phase analysis revealed malaria incidence lagged rainfall by approximately one month and temperature by approximately three to four months, consistent with climate-modulated transmission processes. These findings highlight the heterogeneity of climate-malaria dynamics across transmission settings with contrasting epidemiological implications within Nigeria. The observed lag structures provide a basis for climate-informed early warning systems and intervention timing. While non-climatic drivers were not explicitly modelled, the analysis focuses on isolating climate-driven temporal signals. Consequently, to sustain control and elimination progress, climate-adaptive surveillance and region-specific interventions that anticipate rainfall- and temperature-driven transmission cycles must be integrated into Nigeria’s malaria control framework to ensure timely, targeted, and climate-resilient public health responses.

**Author summary:** Malaria transmission does not respond uniformly to climate drivers across epidemiological settings, highlighting the need to explore climate-malaria dynamics within heterogeneous contexts. Identical climatic forcing can produce qualitatively different outcomes depending on the underlying epidemiological setting, indicating the limitations of generalising control efforts from a single context. Motivated by the need to understand these differences, in this study, we examined the cross-epidemiological scale-dependent and lag-specific climatic forcing of malaria transmission at the sub-national context, providing support for malaria control and elimination strategies. We addressed the following questions to understand the hidden patterns of the temporal cycles and the corresponding associations between the climate variables and malaria incidence in the two states:

1. What are the dominant temporal cycles in malaria incidence in the study region?
2. How do the periodicities of climate variables compare with those of malaria incidence?
3. Are there significant time-dependent associations between climate variability and malaria incidence?
4. How do these association vary across different time scales (intra-annual vs interannual) and periods?
5. What is the average lag between changes in key climate variables and malaria incidence?

Monthly malaria incidence data and corresponding rainfall and temperature records spanning 2015–2024 were analysed using a continuous wavelet transform (CWT) framework. Scale-specific periodicities were identified using wavelet power spectra, while climate–malaria associations were quantified using cross-wavelet power and wavelet transform coherence (WTC). Phase difference analysis was employed to characterise time-varying lead–lag relationships between malaria incidence and climatic drivers at the annual timescale.

Results show that in Lagos, malaria incidence is irregular and weakly linked to climate, reflecting the impact of interventions and socio-environmental factors that disrupt transmission. In contrast, Zamfara exhibits strong, regular annual cycles tightly coupled to rainfall and temperature, with malaria incidence lagging rainfall by about one month and temperature by three to four months.

These findings highlight the need for region-specific strategies: sustaining intervention-driven disruption in low-burden urban areas, and intensifying climate-adaptive measures in high-burden rural settings. Integrating climate-sensitive surveillance and tailored intervention timing into Nigeria’s malaria control framework will strengthen resilience and accelerate progress toward elimination.

Specifically, our findings demonstrate evidence-based framework to guide climate-adaptive intervention timing. In Zamfara state, extreme heat between March and May as shown in the temperature profile, may reduce use of LLINs, indicating that mass distribution before and during these periods, within same year, may be less effective. The start of rain comes with a cooling effect which may facilitates good weather condition that encourages LLIN utilization. Correspondingly, LLIN distribution campaigns conducted in June or July, prior to peak rainfall and peak malaria incidence typically observed between August and October, may enhance intervention effectiveness. Coupled with other climate-sensitive control interventions (for example, seasonal malaria chemo-prevention), such campaigns should be repeated at intervals of no more than three years, in alignment with the observed multi-annual cycles of malaria incidence, to effectively mask malaria risk in Zamafara state. This implementation strategy could be employed in other high transmission states of Nigeria to mitigate malaria risk.

## Introduction

Malaria is a deadly infectious disease caused by protozoan parasites of the genus *Plasmodium*, transmitted through the bites of infected blood-seeking female *Anopheles* mosquitoes [1]. Malaria continues to threaten lives in Nigeria with about 97% of the population estimated to be at the risk of malaria [2]. The heterogeneous dynamics of malaria transmission across different regions driven by diverse climatic patterns makes a “one-size-fit-all” approach inadequate to mitigate malaria transmission. With growing concerns on how climate variability and change may impact vector-borne diseases [3] -of which malaria remains the most significant - objective and evidence-based decision-making are crucial to achieve control and elimination of malaria at different burden zones. The burden of malaria in Nigeria from the global context underscores the urgency of such effort as highlighted in recent reports.

The World Health Organisation Malaria Report 2024 highlighted that Nigeria have the largest share of the global malaria burden [4]. Of the global cases and deaths due to malaria, Nigeria contributed 25.9% and 30.9% respectively, making the country the most malaria-endemic country in the world [4]. Despite high burden and the challenges of COVID-19 pandemic, malaria incidence fell by 26% since 2000, from 413 per 1000 population in 2000 to 306 per 1000 population in 2021 [5]. Ere the pandemic in 2020, malaria was 302 per 1000 population [5]. Leading interventions that contributed to the the decrease in morbidity and mortality rates due to malaria include the scale-up use of long-lasting insecticidal nets (LLINs) for vector control, widespread acceptability and use of artemisinin-based combination therapy (ACT) to treat uncomplicated malaria which rose from 38% in 2015 to 72% in 2021 among children under age five leading to the reduction of malaria among the U5 decreased from 42% in 2010 to 22% in 2021, improved diagnostics, the coverage for the intermittent preventive treatment of malaria in pregnancy (IPTp) - receiving two or more doses of sulfadoxine-pyrimethamine (SP) - which rose from 22% in 2015 to 26% in 2020 [5], seasonal malaria chemo-prevention (SMC) in children under the age of 5 years expanded to 18 states covering about 24 million children [2].

However, while these achievements demonstrate significant progress, the persistence of malaria necessitates a comprehensive and forward-looking strategy to sustain and accelerate gains. The climatic conditions of Nigeria provide suitable environment for the sustainability of the mosquito population thus contributing to the burden of malaria among several other factors [2]. Rainfall and temperature are part of the major climate variables contributing to the conditions suitable for malaria transmission across various transmission settings. Rainfall provides the breeding sites for mosquito development and oviposition while temperature affects the survivability, development and biting activities of the mosquitoes [6]. The different patterns of these climatic variables and their relationship with malaria incidence in different transmission settings create a complex dynamics that translates to heterogeneous epidemiological implications.

Numerous studies have examined the impact of climate variability and change on malaria [6–10]. However, most of these studies have been conducted at broad national or global scales, without evaluating sub-national differences. While such studies provide valuable general insights, their findings may not adequately inform state-level strategies for malaria control and elimination. Consequently, efforts to appropriately guide sub-national strategies towards control and elimination of malaria require studies that involve analyses of the relationship between climate variables and malaria incidence at the sub-national scale. The results of such localised analyses could provide evidence-based guidance that supports tailored, and context-specific strategies to facilitate malaria control and elimination across different regions in Nigeria.

The deployment of control interventions in a malaria-endemic region, like Nigeria, with heterogeneous transmission patterns requires proper guidance on timing to mitigate the intensity of malaria transmission. The dynamic behaviour of epidemiological and climatological processes and the association between them are being identified using statistical framework such as time-series analysis [11, 12]. However, the time-series approach is less reliable for non-stationary epidemiological datasets [13]. The complexity, randomness and non-stationarity of epidemiological time-series require tools beyond the classical techniques that only cater for stationary data. Time series approaches such as ARIMA and its variant models statistically require that the data be stationary. Alternative methods like Fourier analysis require predetermined scaling which may not be suitable for epidemiological time-series with a wide range of dominant frequencies [14]. Thus, a framework of time-frequency that is scale-independent and does not assume stationary data, such as wavelet transform approach should be employed.

Wavelet analysis does time-frequency analysis of epidemiological data to identify and understand patterns, such as seasonality and synchrony between time series that may not be stationary [15, 16]. It provides information on how the frequency characteristics of a disease’s spread change over time, which is particularly useful for studying phenomena like influenza, COVID-19, or changes in seasonality over long periods [17]. This method provides both time and scale information, offering a more detailed view than traditional methods, and can also be used to compare multiple time series to understand their relationships, such as the link between climate and climate-sensitive vector-borne diseases like malaria. Wavelet analysis provides a framework for improving our understanding of the dynamic association between climate variables and malaria incidence [13, 18, 19]. This is of great importance to design and implement effective intervention and control strategies to mitigate malaria transmission across burden zones as related studies have shown.

Oniyelu *et al*., [20] examined variability in malaria incidence among pregnant women compared to general malaria cases across six Nigerian states, using time–frequency decomposition to identify transmission cycles and lead–lag relationships for guiding intervention timing. However, the study did not account for the climatic drivers of the dominant annual and intra-annual cycles, thereby limiting its potential to inform climate-based intervention strategies. Bakare *et al*., [21] analysed the monthly incidence of monkeypox in Nigeria between January 2021 to October 2024 using the wavelet transform approach. The local wavelet power spectrum showed the presence of a strong annual periodicity thus uncovering the hidden patterns of mpox outbreak in Nigeria. Mahendran *et al*., [13] applied wavelet analysis to assess how environmental variability influences malaria transmission in a rural dry-zone locality of Sri Lanka. Significant intra-annual and inter-annual cycles of malaria incidence highlighted the role of climatic fluctuations in shaping transmission dynamics. These studies outline the advantages of using wavelet transform approach for epidemiological data to assess variations of temporal cycles and how key drivers influences them.

Notwithstanding, little or no studies have been conducted to examine the variations in temporal cycles of climate variables and their associations with malaria transmission in Nigeria. Lagos and Zamfara, where the current study was carried out, are two States representing malaria-endemics regions with different epidemiological settings. These two states have contrasting transmission intensities, human settlement and ecological characteristics. Several studies have been conducted in these states establishing the relationship between malaria and various environmental and socio-economic factors. These studies include identification of the relationship between malaria prevalence and environmental factors [22], assessment of the effect of malaria knowledge, attitude, perception and practices on malaria prevalence [23] and the role of poverty and economic situation on malaria prevalence [24].

However, there is paucity of studies that address variations in temporal cycles of malaria incidence and its association with climatic drivers in different epidemiological settings. Consequently, this creates a crucial gap in the knowledge required to guide decision-making towards control and elimination of malaria in Nigeria. The aim of this study is to address this gap by examining the scale-dependent and lag-specific climatic forcing of malaria under heterogeneous transmission settings.

## Materials and methods

### Study Setting

Lagos and Zamfara are two states among the 36 states in Nigeria located at different ecological and malaria-burden settings. Lagos state is located in the South-West Nigeria with a projected population of 16.5 million in 2024 [25]. Lagos state has a humid tropical climate with distinct wet and dry seasons that runs from March to October, and November to February respectively [5, 26]. Maximum temperature ranges between 29^*o*^ C and 34^*o*^ C, and minimum temperature varies between 24^*o*^ C and 28^*o*^ C with mean annual rainfall averaging around 1535.8 millimeters [5, 26]. Of the Nigeria’s 68 million malaria cases reported in 2021, Lagos state contributed an estimated 3.8%. Malaria prevalence in Lagos state reduced from 15% in 2010 to 2.6% in 2025, the lowest in Nigeria which makes Lagos state a low transmission setting [5, 27]. The location of Lagos is delineated on map of Nigeria shown in Fig 1.

**Fig 1.**
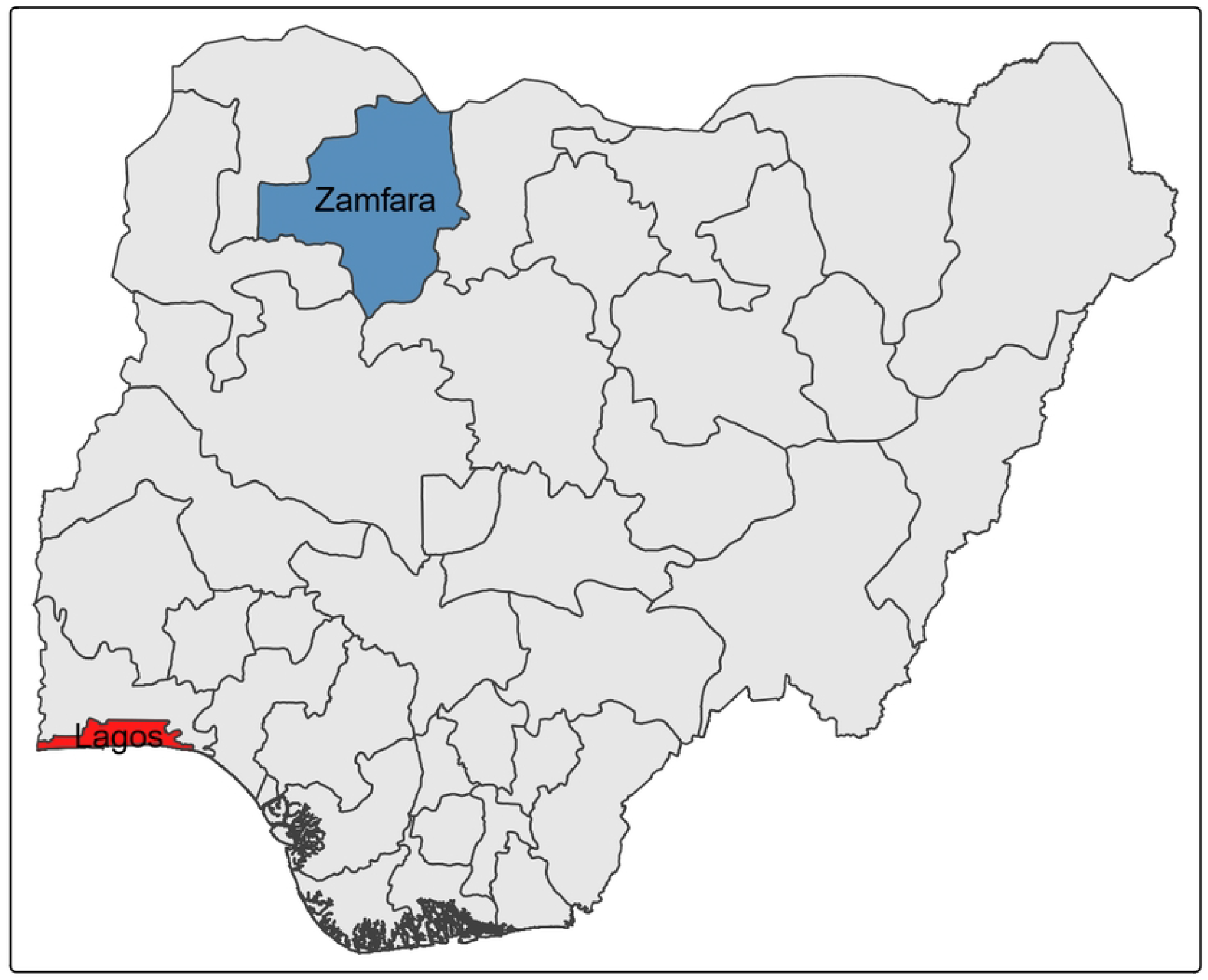
Map of Nigeria showing the study states. The highlighted states represent the low- and high-malaria-burden regions included in this study.

Zamfara is located in the North-West Nigeria with a projected population of 5.4 million in 2022 [5]. Ecologically located at the Sudano-Sahelian zone, the state is characterised by two distinct seasons: the wet season (May to September) and the dry season (October to April) [5]. This leaves a window of 7 months for dry season in the state. The climate of Zamfara is tropical with a temperature rising up to 38^*o*^ and above between March to May [28]. Second to Kebbi (the state with the highest malaria prevalence of 52% by microscopy), Zamfara state has a prevalence of 36% due to malaria which ranks the state among the high-burden states in Nigeria [5].

### Data Description

#### Monthly Uncomplicated Malaria Data

We used the monthly uncomplicated malaria incidence data of Lagos and Zamfara states which span January 2015 to December 2024 obtained from the national malaria data repository (NMDR) to conduct the analysis. The time series plots of the monthly malaria incidence data for the two two states are presented in Fig 2(a) and (b).

**Fig 2.**
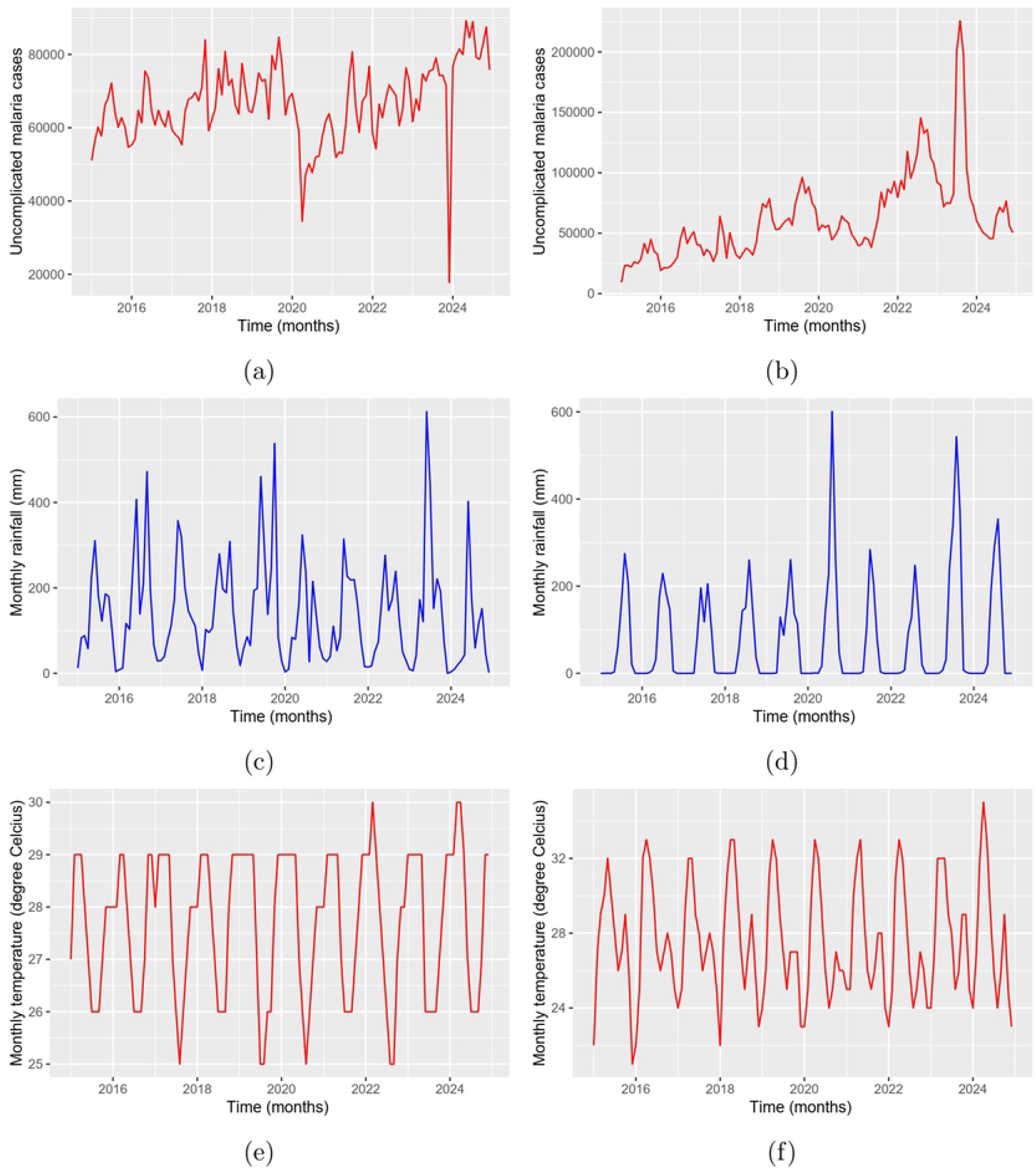
Time series of malaria cases and climatic variables in Lagos and Zamfara States. Monthly trends of uncomplicated malaria cases, rainfall, and temperature in Lagos State (a, c, e) and Zamfara State (b, d, f).

#### Climate Data

Monthly rainfall and temperature data for both states were obtained from the World Weather Online Database (WWOD) [29]. Climatic data from the state capitals were used as proxies for state-level conditions, consistent with existing climate–malaria modelling studies.

### Wavelet Transform Analysis

In this study, we use wavelet analysis, a framework widely applied in various fields including infectious diseases such as monkeypox [21], malaria [20], influenza [30], and COVID-19 [31]. Wavelet transform approach reveals the time-frequency domain for statistically non-stationary epidemiological and climatological time series, and also highlights both positive and negative association simultaneously [30]. We adopted the continuous wavelet power spectrum, and three cross-wavelet tools: cross-wavelet power spectrum, wavelet coherency and wavelet phase difference in the WaveletComp library (version 1.2) implemented in R.

#### Wavelet Transform

The continuous wavelet transform is applied in this study to investigate the seasonal fluctuations of malaria incidence, rainfall and temperature in the locations with different transmission intensity. Given a single time series *x*(*t*) in a continuous time, *t*, the basic wavelet transform is a function of the time domain and frequency (expressed inversely by the scale or period, *s*). It is mathematically defined as

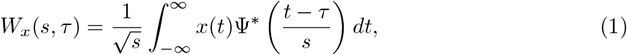

where Ψ^∗^(.) is the complex conjugate of the mother wavelet which is shifted in time by *τ* and scaled by *s* [15].

#### Morlet Wavelet

The WaveletComp (version) analyses frequency structure of uni- and bivariate time-series using the Morlet wavelet [32]. The mother Morlet wavelet implemented in WaveletComp is:

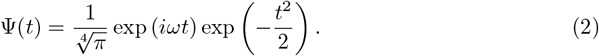

The angular frequency, *ω*, is set to 6, which is the preferred value in literature since it makes the Morlet wavelet approximately analytic [14, 32].

#### Amplitude and Wavelet Power Spectrum

The amplitude and wavelet power spectrum given respectively as

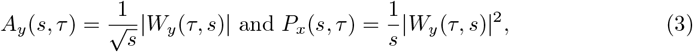

are concepts derived from the basic wavelet. The power spectrum is simply the square of the amplitude. The importance of periodic fluctuations of scale, *s* (period) at a time *τ* is reflected by the magnitude of the wavelet power spectrum.

#### Wavelet Coherency and Phase Difference

Continuous wavelet transform provides the time-frequency domain, where frequency changes over time using the base Morlet Mother wavelet function. To guide policy-making regarding the mitigation of malaria transmission, it is essential to quantify the relationship between malaria incidence and each of the climate variables. The wavelet transform coherence is applied to investigate the coherency between two series, X and Y. The wavelet cross-spectrum and wavelet coherence can be computed to quantify the relationship between two non-stationary signals [15]. The wavelet cross-spectrum is given by

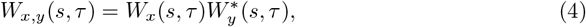

where denoted the complex conjugate.

The wavelet coherency is defined as the cross-spectrum normalized by the spectrum of each signal,

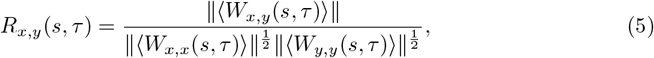

where “ ⟨⟩” denotes a smoothing operator in both time and scale [15].

The phase difference between two epidemiological signals *x*(*t*) and *y*(*t*) is mathematically defined as:

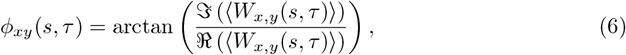

to characterize the possible relationship between the two signals at specified timescale [15].

### Analysis of data

In this study, the wavelet analysis was performed using wavelet toolbox of the WaveletComp library implemented in the R software. The data were detrended to remove the underlying low-frequency trend from the time series to focus on the oscillations and periodicities. The continuous wavelet transform, average wavelet power spectrum, and cross-wavelet spectrum were generated for the monthly time-series data of malaria incidence and climatic variables assuming a 5% level of significance. Areas of statistically significant power are indicated by the white contour lines, with red regions inside these contours denoting strong wavelet power, green representing moderate power, and blue signifying weak wavelet power. Statistical significance was assessed using Monte Carlo simulations (n = 1000) under a red-noise null hypothesis, with regions exceeding the 95th percentile considered significant.

The wavelet power spectra were used to identify dominant periodicities in malaria incidence and climate variables, as well as their temporal variations. Cross-wavelet power spectra and wavelet coherence were used to examine the strength and stability of associations between the climatological and epidemiological data across time. Phase difference analysis was applied to characterise the time-dependent lead-lag relationship between the climate variables and malaria incidence, with particular focus on lag-specific variations at the annual timescale.

## Results

### Continuous wavelet transform of malaria incidence and climate variables

The continuous wavelet transforms (CWTs) of malaria incidence and the climate variables for Lagos and Zamfara states are shown in Fig 3 (Lagos) and 4 (Zamfara). To identify and characterize the temporal cycles of climate variables and malaria incidence, local wavelet power spectrum analysis was carried out on the respective time series data for Zamfara and Lagos States. In Fig 3, we presented the local wavelet power spectrum of the monthly malaria incidence, temperature, and rainfall for Lagos State. In Fig 3 (a), monthly malaria incidence in Lagos State shows intermittent cycles ranging from semi-annual (6-month), annual (12-month), and multi-annual (24- and 36-month) cycles that are transient, reflecting a disruption in the seasonality of the malaria incidence.

**Fig 3.**
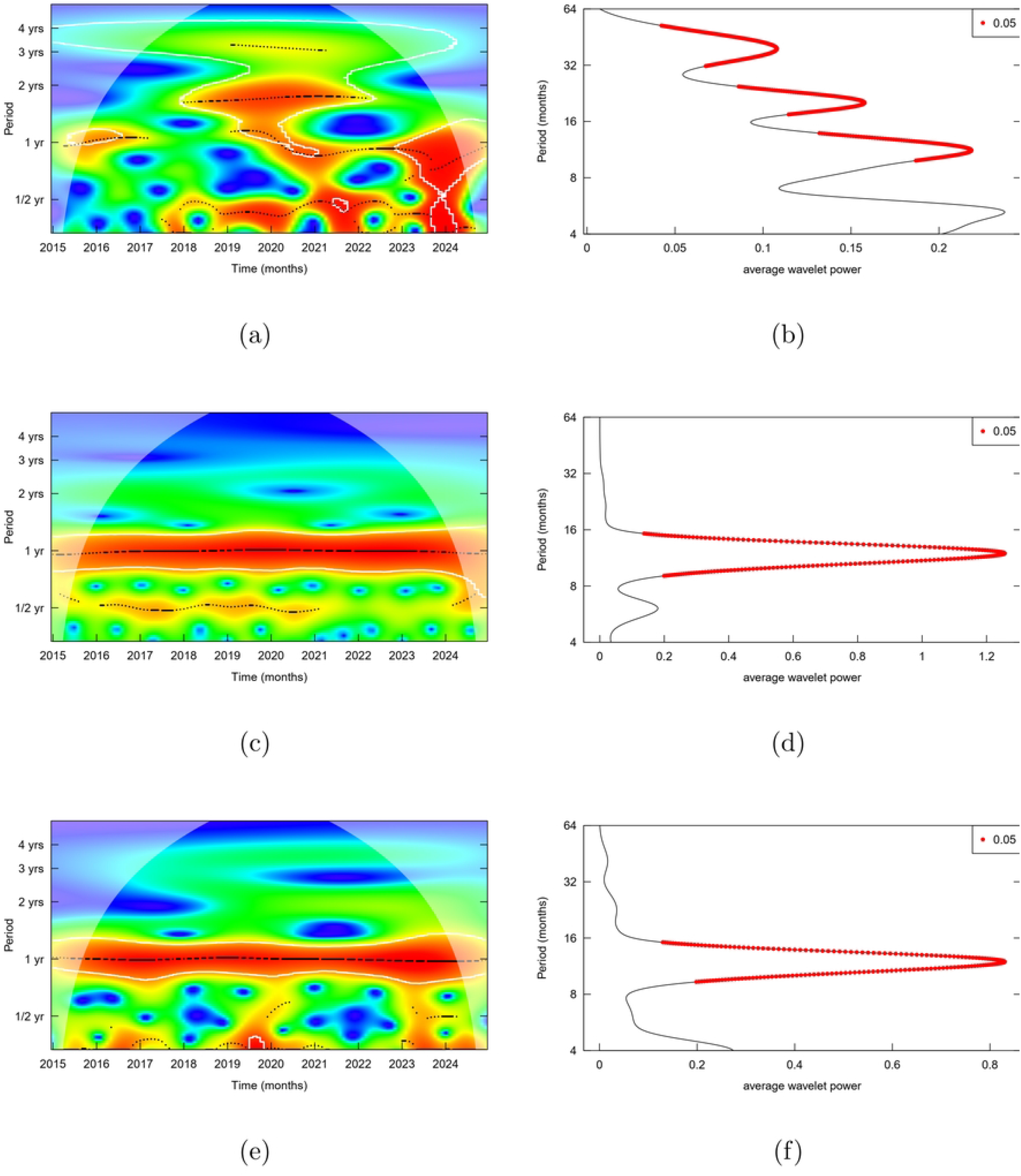
Wavelet power spectra and local wavelet spectra of climatic variables and malaria incidence in Lagos State (2015–2024). Wavelet power spectrum and corresponding local (time-averaged) wavelet spectrum of monthly (a) malaria incidence, (b) temperature, and (c) rainfall.

The local wavelet power spectrum of the temperature reveals a persistent dominant annual cycle as shown in Fig 3 (b). This reflects the environmental conditions of the dry and wet seasons in Lagos State, as cooling of the environment coincides with the start of rainfall. The rainfall patterns similarly exhibits strong and significant dominant annual cycle which reflects the condition of rainfall in Lagos State, consistent with the tropical rainforest ecological zone in which Lagos is situated. The cone of influence, delineated by the lighter shaded area outlines the region where edge effects are significant. The areas of highest power (red and yellow) are outside this cone, confirming that these cycles are not artifacts of the data boundaries.

The monthly malaria incidence in Zamfara exhibits a significant dominant annual cycle as shown in Fig 4 (a) throughout the study period. Statistically significant regions, identified at the 5% level, are concentrated around the 12-month band, indicating strong and regular seasonality in malaria transmission. Aside the dominant annual cycle, there are significant intermittent intra-annual and multi-annual cycles reflecting intra-annual and multi-annual variability of malaria transmission in Zamfara. The consistent presence of significant power at the annual scale suggests that malaria incidence in Zamfara is likely modulated by seasonal climatic factors, with minimal disruption from non-climatic influences.

**Fig 4.**
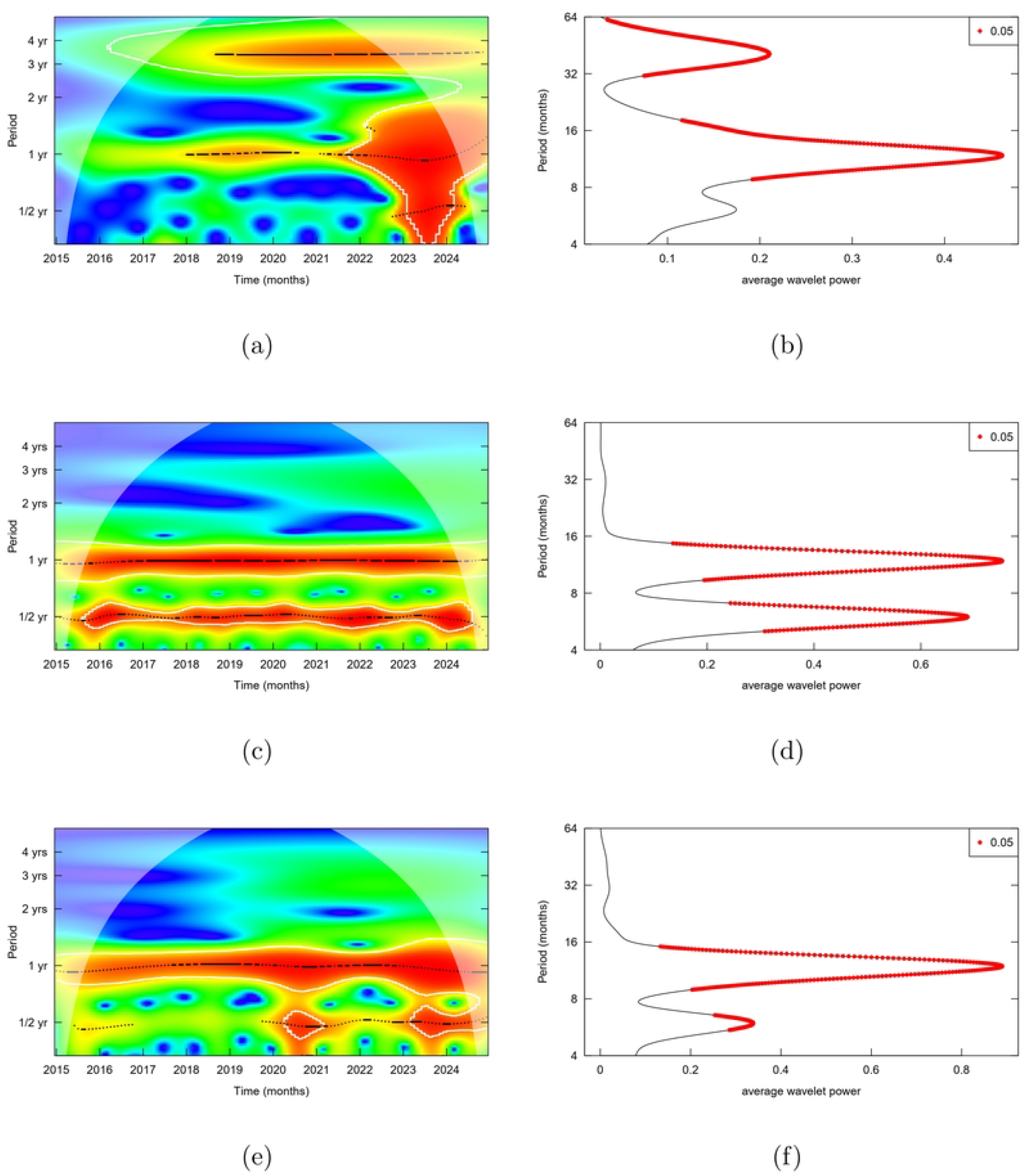
Wavelet power spectra and local wavelet spectra of climatic variables and malaria incidence in Zamfara State (2015–2024). Wavelet power spectrum and corresponding local (time-averaged) wavelet spectrum of monthly (a) malaria incidence, (b) temperature, and (c) rainfall.

In Fig 4 (b), the local wavelet power spectrum (LWPS) of the monthly temperature shows a strong and significant annual and semi-annual periodicities over time. Statistically significant semi-annual periodicity is shown in Fig 4 (a) and (b) for malaria incidence, and temperature signifying association with malaria incidence. Similarly, Fig 4 (c) reveals a strong annual seasonal peaks of rainfall in Zamfara over time which is likely responsible for the notable annual seasonal pattern in the malaria incidence as shown by local wavelet power spectrum.

Furthermore, a remarkable semi-annual seasonal pattern of rainfall emerged at the middle of 2020 until 2024 which run concurrently with the annual rainfall seasonal pattern pointing to a change in the rainfall pattern in Zamfara state. This change in the rainfall patterns has the potential to contribute to the availability of more breeding sites depending on the hydrological characteristics of Zamafara state. A close observation of the rainfall and malaria incidence patterns in Zamfara state towards the end of 2024 from the middle of 2023 through the LWPS reveals semi-annual seasonal patterns in the monthly rainfall and monthly malaria incidence in Zamfara state. These observed bimodal cycles of the rainfall, temperature and malaria incidence result into two distinct seasonal peaks of malaria transmission per year, leading to a higher overall disease burden and more complex control strategies.

### Cross-Wavelet and Coherence Analyses

Wavelet coherency examines the time-frequency relationships between two time series by identifying localised correlations in both time and frequency domains. The association between the temporal cycles of the malaria incidence and climate variables is demonstrated using wavelet transform coherence (WTC) as shown in Fig 5 and 6. The coherence between temperature and malaria incidence of Lagos state is displayed in Fig 5 (a). The area within the white contour lines shows the region of significant periods for the WTC between temperature and malaria incidence and the band of colours (blue → green → yellow → red) signify their common strength as shown by the scale bar at the right hand side of the graph.

**Fig 5.**
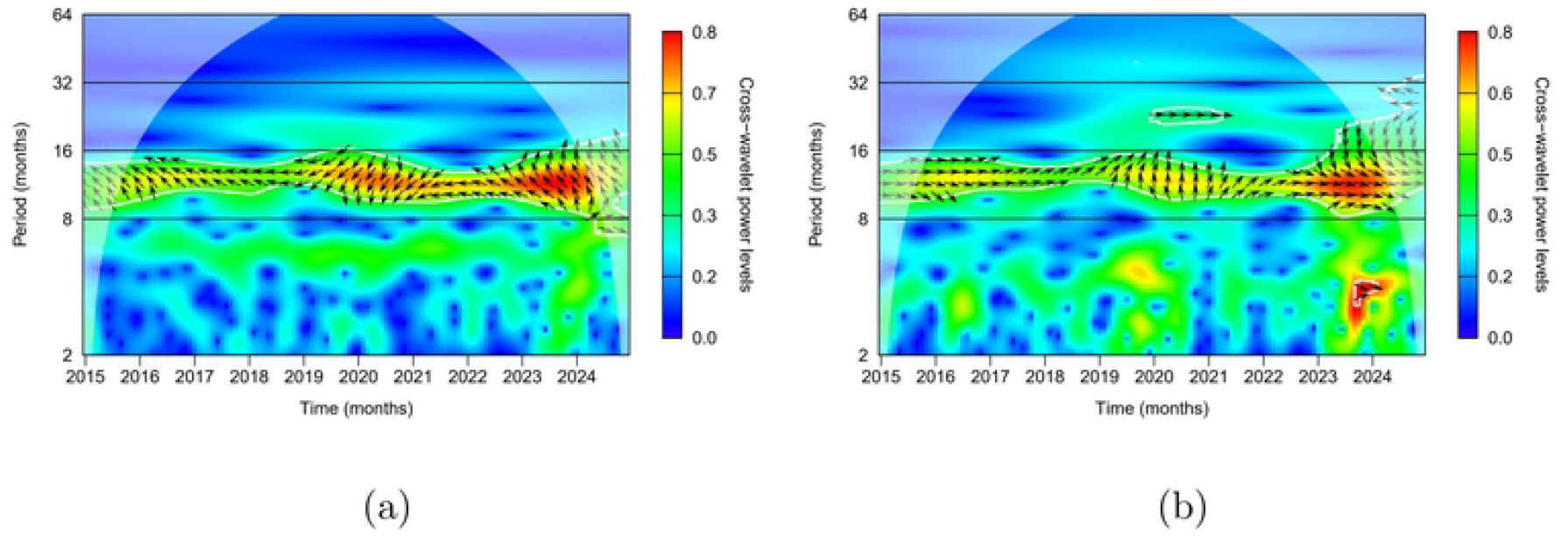
Cross-wavelet transform between climatic variables and malaria incidence in Lagos State (low-burden urban setting). Cross-wavelet transform of (a) temperature and (b) rainfall with malaria incidence. Color intensity represents the magnitude of the common power in the time–period domain, with red (blue) indicating high (low) power. The thick white contour denotes regions significant at the 5% level against a red-noise background. The shaded region indicates the cone of influence (COI), where edge effects may distort results. Arrows represent the phase difference between malaria incidence and the climate variable: arrows pointing right (left) indicate in-phase (anti-phase) relationships; arrows pointing right-up (right-down) indicate the climate variable leading (lagging); arrows pointing left-up (left-down) indicate leading (lagging) relationships under anti-phase conditions.

**Fig 6.**
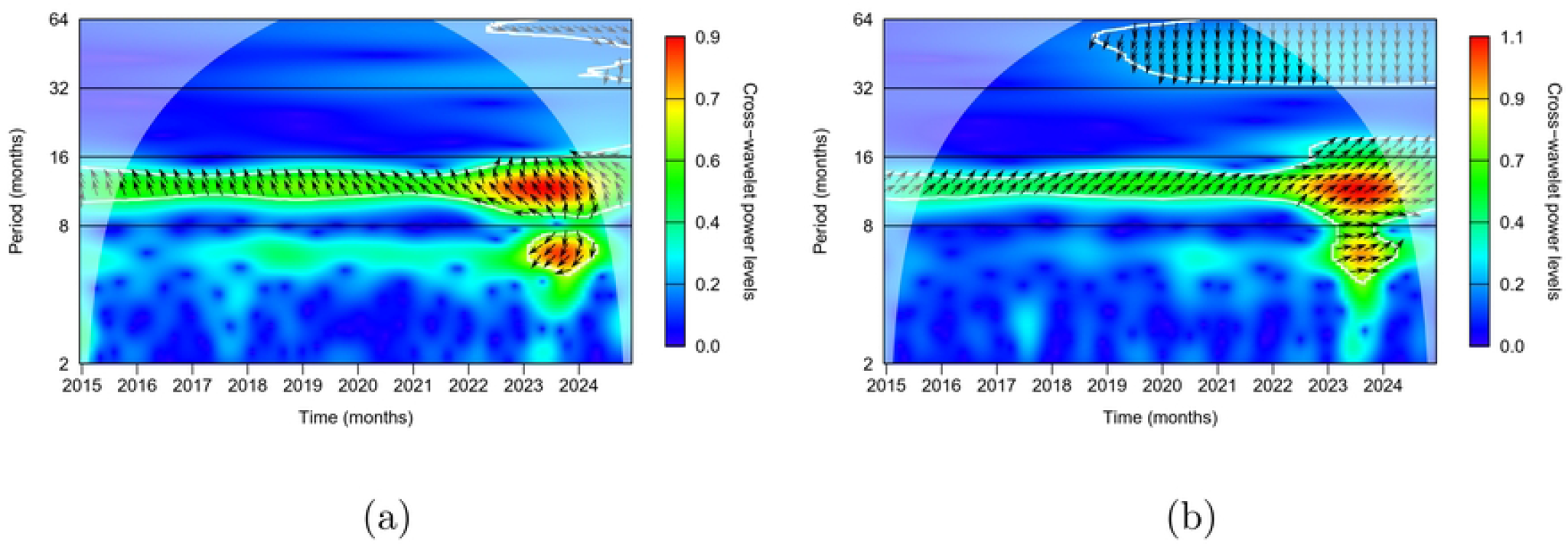
Intra-state cross-wavelet analysis in Zamfara State. Cross-wavelet coherence between malaria incidence and (a) temperature, and (b) rainfall in Zamfara State (a high-burden, predominantly rural state). Warmer colours (red) indicate stronger coherence, while cooler colours (blue) indicate weaker coherence in the time–period domain. The colour scale is shown on the right-hand side of each panel. The thick white contour denotes the 5% significance level against red noise. The shaded region represents the cone of influence (COI), where edge effects may distort results. Arrows indicate phase differences: right (left) arrows denote in-phase (anti-phase) relationships; arrows pointing upward (downward) indicate that the climate variable leads (lags) malaria incidence.

Approximately, from 2016 to 2024, the dominant cycles appear within the 8-16-month band of the period (highlighted in green → yellow → red). This indicates that temperature and malaria in Lagos State share a common cyclical or seasonal patterns that approximately correspond to annual to 1.5-year cycles as outlined in Fig 5 (a). The red/yellow regions within the 8-16-month band reflect periods where the climatic forcing is dominant. Specifically, a strong dominant annual cycle between the temperature and malaria incidence is highlighted from 2019 to 2023 in Lagos State, as indicated by the red/yellow colour band. The cone of influence (COI) indicates the region where edge effects are minimal. Most of the strong 8–16-month band lies within the COI, confirming that the results are statistically robust and reliable.

Additionally, Fig 5(a) presents the wavelet transform coherence between temperature and malaria incidence in Lagos State. The thick white contour outlines regions of statistically significant coherence, which largely coincide with the 8–16-month band where power is strongest. This indicates a consistent and significant relationship between temperature and malaria incidence across time within this frequency range. The arrows demonstrate the relative phase between the two variables, providing insight into temporal modulation and lagged responses. Their directions vary across the study period: from 2016 to 2018, arrows point diagonally upward to the left, suggesting that temperature leads malaria incidence, although the two are out of phase. Between 2019 and 2021, arrows point downward, indicating that malaria incidence leads temperature. Moreover, from 2022 to 2023, arrows again point diagonally upward to the left, showing that temperature leads malaria incidence with a lag of approximately 1–4 months. These findings suggest that temperature influences malaria incidence with a delayed effect, which may not solely reflect biological development traits of the malaria vector or parasite, but rather time-dependent variations in suitable conditions for transmission intensity. This suggests that control interventions must anticipate future transmission rather than respond only to current case trends.

Fig 5 (b) presents the wavelet transform coherence between rainfall and malaria incidence in Lagos State. From mid-2015 to late 2018, the arrows point eastward, indicating a strong temporal association between the two variables. During this interval, both rainfall and malaria incidence display dominant annual cycles, with the eastward orientation of the arrows signifying that they are in phase—reaching their peaks almost simultaneously. This synchrony highlights the close coupling between rainfall patterns and malaria incidence in Lagos State. In contrast, between 2019 and 2021, the arrows shift northward, showing that malaria incidence lags rainfall by approximately 2–3 months. This delay may likely corresponds to the time required for rainfall to generate stable pools that serve as mosquito breeding sites, followed by the subsequent growth of mosquito populations sufficient to intensify malaria transmission. Notably, while malaria incidence leads temperature during this same period, the coherence analysis demonstrates that rainfall most likely modulated malaria incidence between 2019 and 2021.

Taken together, these findings underscore the critical role of rainfall in shaping malaria transmission dynamics and highlight the importance of integrating climate-based surveillance into malaria control programs in Lagos State. By anticipating rainfall-driven transmission peaks, Lagos State’s public health authorities can better time interventions such as vector control, community sensitization, and distribution of preventive measures to reduce the burden of malaria in Lagos State which has the potential to take the State to pre-elimination stage.

Fig 6 demonstrates the cross-wavelet power spectrum and wavelet transform coherence between the climate variables and malaria incidence in Zamfara State. Similar to the dynamics of rainfall and temperature in Lagos state, the significant region of the cross-wavelet power spectrum lies between the 8-16-month band delineated by the thick white contour line. This reflects common temporal cycles, a roughly annual to multi-annul cycles with moderate to strong power, shared between the climate variables and malaria incidence in Zamfara (a high-burden malaria-endemic State).

Besides the common temporal cycles shared between the climate variables and malaria incidence in Zamfara, Fig 6 (b) shows a significant biennial relationship between rainfall and malaria incidence outlined by the white contour line within the 16-32-month band of the cross-wavelet power spectrum. Remarkably, a statistically significant semi-annual temporal cycles variation is also observed between the rainfall and malaria incidence below the 8-month boundary of the cross-wavelet power spectrum indicating a change in the pattern of malaria likely influenced by the shift in rainfall pattern during the 2023 to 2024 interval.

The wavelet transform coherence analysis shows a consistent temporal relationship between the climate variables and malaria incidence in Zamfara. Fig 6 (a) shows arrows that consistently diagonally point up to the left, that is, northwest, highlighting that temperature continuously leads malaria incidence throughout the observed period though both are out of phase. This reflects a very weak, or no synchrony between the malaria incidence and temperature in Zamfara State. Furthermore, within the year 2023, a significant semi-annual seasonal forcing between temperature and malaria incidence is seen with the arrows showing that malaria increases before rainfall. While this is biologically unlikely, the co-occurrence of the semi-annual and annual temporal cycles provides hints on the possibility of this scenario. The increase in malaria due to annual suitable temperature conditions sufficient for malaria intensity mostly appears before the semi-annual cycle, whose increase may not be suitable for the intensity of malaria incidence. This unlikely event underscores the need to integrate climate-based analyses with entomological and epidemiological evidence when designing malaria surveillance and control strategies.

Coherence between the rainfall and malaria incidence propagates consistently through the observation period indicating a strong relationship between the two variables in Zamfara State. Fig 6 (b) highlights the arrows within the 8-16-month band of the period pointing diagonally upward towards the right. The signifies a reflection of the lead-lag relationship between the two variables. Specifically, the arrows dominant within the annual seasonal band shows that increase in rainfall consistently lead malaria incidence by approximately 1-2 months. Unlike the irregular patterns observed between the rainfall and malaria incidence in Lagos State, which may be attributed to disruption of climate forcing due to control measures against the intensity of malaria transmission, the regular climatic influence on malaria incidence in Zamfara State underscores the need to intensify effort to disrupt the influence of climatic factors on malaria transmission.

### Phase Analysis of Climate Variables and Malaria Incidence

The phase analysis of the wavelet transform coherence illustrates the method to quantify the lead-lag relationship between the climate variables and malaria incidence. Dominant 12-month cycles confirm that malaria transmission is strongly and consistently driven by predictable seasonal factors like rainfall and temperature. This allows for the effective planning and timing of annual control measures, such as indoor residual spraying, seasonal malaria chemo-prevention and bed net distribution, to coincide with the high-transmission season. Consequently, for policy and decision-making relevance, the phase analysis is performed considering only the annual timescale to recommend climate-sensitive intervention timing, thereby guiding efforts towards control and elimination of malaria transmission across states with varying burden level.

Fig 7 and 8 show the phase relationship between the climate variables and malaria incidence in Lagos and Zamfare states. Fig 7 (a) shows the phase angle for the annual component of the wavelet transform coherence between temperature and malaria incidence in Lagos State. The phase analysis reveals an irregular synchrony between the two variables over the studied period. Specifically, the temperature shows an irregular relationship between 2019 and 2021, with malaria incidence leading the temperature, suggesting that rainfall was more influential during this window period, as revealed by the coherence analysis. The phase diagram further shows that malaria incidence increases as the temperature declines from the peak signifying that temperature and malaria has an indirect relationship.

**Fig 7.**
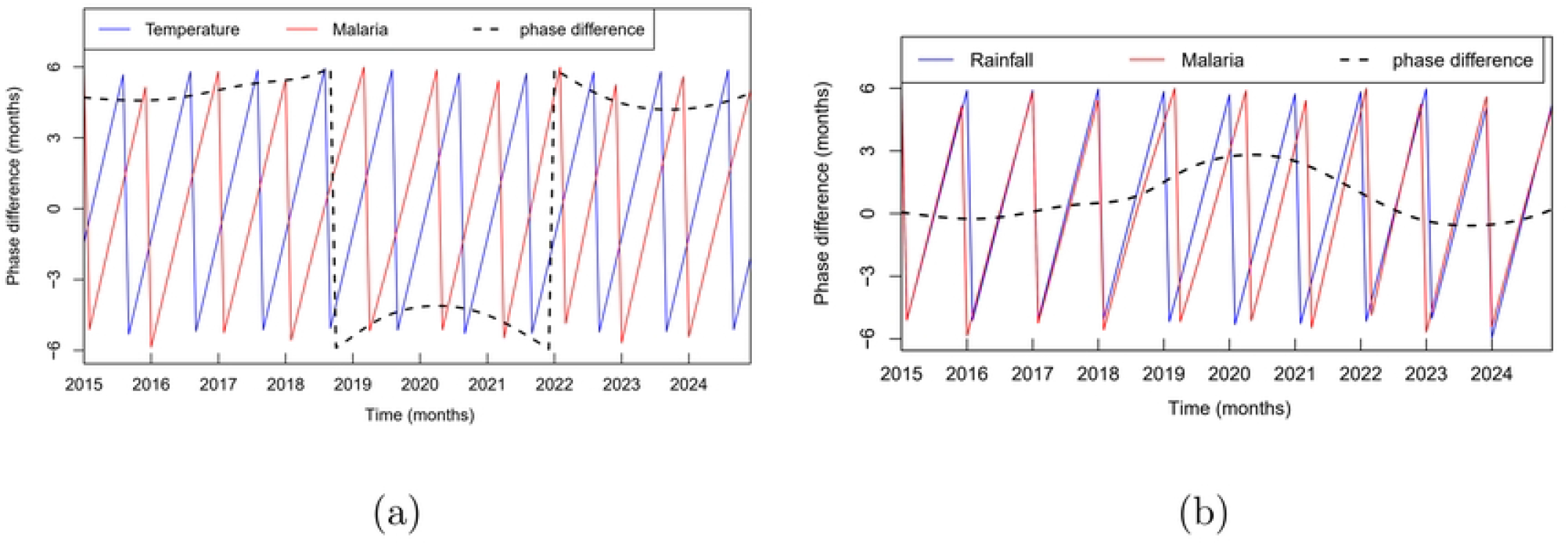
Phase difference analysis. Phase relationships showing time-dependent lead–lag associations between malaria incidence and climate variables: (a) temperature and (b) rainfall in Lagos State, 2015–2024.

**Fig 8.**
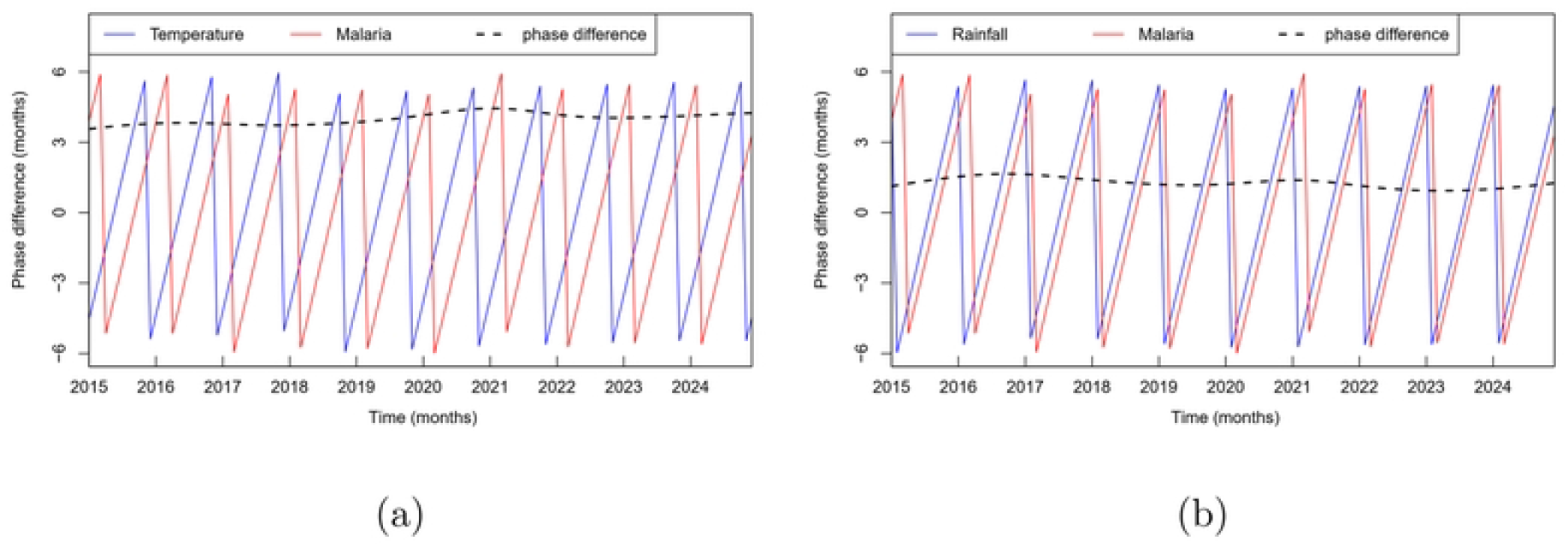
Phase difference analysis. Phase relationships illustrating time-dependent lead–lag associations between malaria incidence and climate variables: (a) temperature and (b) rainfall in Zamfara state, 2015–2024.

However, in contrast, the phase analysis highlights a consistent and stable synchrony between rainfall and malaria incidence as shown in 7(b). Precisely, rainfall and malaria shows a strong synchrony between 2015 and 2017, and 2022 and 2024. A lead-lag relationship of approximately one month, on average, between rainfall and malaria within the window period of 2018 and 2021 is highlighted in 7 (b) where rainfall consistently leads malaria incidence. This pattern underscores the dominant role of rainfall in shaping malaria transmission dynamics in Lagos State, particularly during periods when temperature exhibits irregular associations.

The phase analysis of the wavelet transform coherence between climate variables and malaria incidence revealed notable results in Zamfara State. Temperature and rainfall are not synchronous with malaria incidence; rather, both climate variables exhibit a clear and stable lead–lag relationship. As highlighted in Fig 8, rainfall and temperature consistently lead malaria incidence throughout the study period. The annual peaks of rainfall precede the seasonal peaks of malaria incidence by an average of approximately one month, representing a measurable lag between rainfall and malaria transmission in Zamfara. In the same vein, the analysis further highlights that temperature leads malaria incidence by approximately four months on average. These lags reflects biological delays inherent in the transmission cycle and ecological constraints that may hamper vector growth sufficient to cause alarming malaria incidence.

### Wavelet-based spatio-temporal analysis of the variables in the two states

We demonstrated the time-frequency association between malaria incidence, and the climate variables in the two states to empirically assess the variations between them. Fig 9 reveals cross-wavelet power of the malaria incidence between the two states (Lagos and Zamfara). Statistically significant association of malaria incidence between the two states is only observed from 2022 to late 2024 within the 8-16-month band of the power spectrum. Statistical significant association observed between the malaria incidence in the two states remarkably reveals changing patterns in the dynamics of malaria in Lagos state when correlated with that in Zamfara on annual timescale (Fig 9(b)). Non-significant association existed in the previous years indicating structurally heterogeneous climate-malaria response across the two settings.

**Fig 9.**
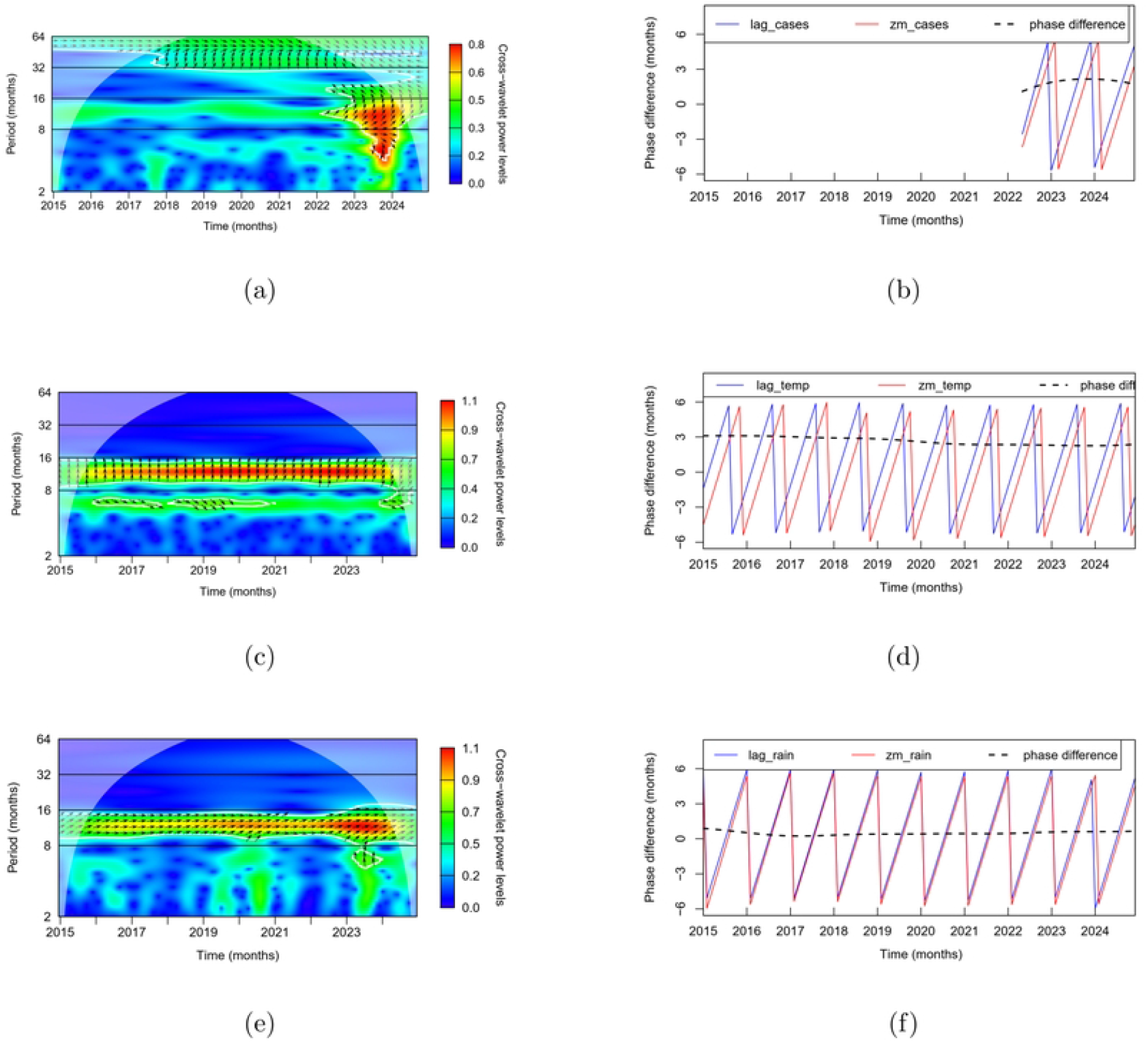
Cross-wavelet analysis of malaria incidence and climate variables in the study states, 2015–2024. Cross-wavelet power spectra and corresponding phase relationships illustrate time–frequency coherence and time-dependent lead–lag associations between malaria incidence and temperature and rainfall. Panels (a), (c), and (e) show cross-wavelet power spectra, while panels (b), (d), and (f) show the associated phase diagrams. All significance levels are tested at 5% against red noise.

The arrows within the 8-16-month band show that there exists a time-dependent lead-lag relationship between the malaria incidence in the two states. Although both states are largely in phase during this period, the arrows show that malaria incidence in Lagos consistently leads that in Zamfara during this significant period. The phase analysis further identifies the phase difference between the two to be approximately two months on average. The phase diagram confirms that the peaks of malaria incidence in Zamfara lag Lagos state by approximately 2 months. These findings highlight heterogeneity in the temporal dynamics of malaria transmission across the two settings.

The climatic variables in the two states also exhibit distinct time-dependent relationships. Temperature in Zamfara consistently lags that in Lagos by approximately three months signifying that temperature reaches its peaks earlier in Lagos on the annual timescale. Similarly, rainfall in Lagos consistently leads that in Zamfara by approximately one month. These spatio-temporal differences in climate signals reinforce the observed heterogeneity in malaria dynamics and underscore the contrasting temporal responses of malaria transmission to climatic drivers across transmission settings.

Pearson correlation analysis of the normalized epidemiological and climatological data was performed to assess intra- and inter-state climate-malaria, and climate-climate linear associations. As shown in Table 1, malaria in both states exhibit weak negative coherence, indicating non-significant linear associations. The results are consistent with the time-dependent localised association as obtained from the wavelet analysis. Conversely, malaria shows moderate and statistically significant correlation with malaria in Lagos state suggesting the linear association and localised synchrony between them. However, malaria in Zamfara exhibits a stronger and significant linear association with rainfall reinforcing the heterogeneous response of malaria to climate variables across settings.

**Table 1.** Pearson intra- and inter-state correlations between malaria incidence and climate variables in Lagos and Zamfara states. Values represent correlation coefficients with 95% confidence intervals in parentheses.

| Pearson intra-state correlation |  |  |
| --- | --- | --- |
|  | Lagos | Zamfara |
| Malaria-Temperature | -0.1517 (95%CI : -0.3222, 0.0282),<br>$p = 0.0981$ | -0.1460 (95%CI : -0.3170, 0.0341),<br>$p = 0.1115$ |
| Malaria-Rainfall | 0.2235 (95%CI : 0.0461, 0.3872), $p < 0.05$ | 0.4150 (95%CI : 0.2547, 0.5531), $p < 0.05$ |
| Pearson inter-state correlation |  |  |
| Malaria-Malaria | 0.2357 (95%CI : 0.0590, 0.3981), $p < 0.05$ | |
| Rainfall-Rainfall | 0.4580 (95%CI : 0.3037, 0.5889), $p < 0.001$ | |
| Temperature-Temperature | 0.3042 (95%CI : 0.1321, 0.4584), $p < 0.001$ | |

Inter-state Pearson correlations between malaria incidence and climate variables in Lagos and Zamfara states reveals a weak significant association despite moderate associations between their climate variables. This observations confirm that, despite shared climatic variability, malaria incidence remains only weakly synchronized across the two states, reinforcing heterogeneous climate–malaria responses.

## Discussion

The investigation of the dynamic patterns of malaria incidence at sub-national scale, considering different epidemiological settings in terms of malaria intensity, urbanisation and climatic zone, requires a comprehensive understanding of its relationship with climatic factors to guide climate-adaptive control and elimination strategies. The analysis performed in this study revealed significant temporal relationship and variation in the periodicities of the climate variables, and how these compare with malaria incidence in low- and high-transmission settings. By translating the observed lead–lag relationships into actionable insights, the analysis provides a framework for optimizing the timing of interventions and strengthening malaria risk mitigation strategies.

Our results highlighted transient but significant semi-annual, annual, and multi-annual cycles of malaria incidence in Lagos State, a low-transmission setting despite consistent annual rainfall and temperature temporal cycles. These inconsistent transient cycles might be attributable to the impact of control interventions deployed to mitigate malaria transmission in Lagos State. The absence of persistent annual periodicity suggests that this disruption is evident among several other factors.

Although control interventions are often cited as drivers of disrupted malaria seasonality, available malaria indicator survey (MIS) and the demography and household survey reports indicate that coverage long-lasting insecticidal nets (LLINs) has remained low and declined over time in Lagos explaining that LLINs alone cannot explain the observed disruption. We suppose that other control measures and anthropogenic activities, not considered in this study, might have played a more decisive role. The expectation that the regular seasonality of the rainfall and temperature should bring about regular seasonality of malaria incidence was contradicted by the local wavelet power spectrum shown in Figure 3(a). This observation portrays complex dynamics that necessitates robust surveillance to mitigate malaria risk in low-endemic regions.

The presence of significant, though transient, annual (12-month), intra-annual (6-month) and multi-annual (18 - 30 months, and 36 months) cycles may imply interruption of the climatic factors due to other factors that may include environmental management that reduces the mosquito breeding sites and survivability, measures that limits contact between human-hosts and malaria vector (window screening with nets, for example) and good drainage system that facilitates washing away of breeding sites that hamper the sustainability of the mosquito population required for intense consistent seasonal transmission of malaria. The presence of significant but transient intra-annual and multi-annual malaria cycles further indicates that malaria transmission in Lagos is characterized by irregular dynamics, highlighting the limitations of assuming fixed seasonal patterns in low-burden settings.

Precisely, presence of intra-annual and multi-annual cycles suggests that a nominal approach may not be sufficient to mitigate malaria transmission in Lagos State. Consequently, flexibility of public health strategies and ability to account for longer-term fluctuations are crucial to mitigate malaria transmission with such complex dynamics observed in Lagos State. Robust surveillance to detect the start of these multi-year trends is very crucial to mask future malaria incidence as inter-annual variability in malaria incidence can affect the assessment of the impact of malaria interventions.

In contrast, climate variables remain major drivers of malaria transmission in Zamfara State. The local wavelet power spectrum revealed significant and regular annual cycles (as shown in Figure 4) highlighting the alignment of malaria incidence with rainfall and temperature variability. These findings reinforce that, unlike Lagos where anthropogenic interventions and activities might have weakened climatic forcing, malaria transmission in Zamfara tends to be strongly climate-driven. This underscores the urgent need to intensify control and elimination efforts aimed at disrupting the influence of rainfall and temperature on malaria incidence in Zamfara. Strengthening climate-adaptive strategies in high-burden states will be critical to reducing transmission intensity and sustaining progress toward malaria control and elimination.

Similar transient multi-annual cycles were recorded in Venezuela, as in Lagos, emphasising the complex dynamics of malaria-climate relationship varying in strength across the studied regions [33]. Malaria was found to exhibit unique features in each of the studied region like the case of the two States in this study. Similarly, malaria cases in Sri Lanka exhibited significant intermittent periodicities in the 6^th^ to 12^th^ month and in the 2^nd^ to 5^th^ month at different window period, and other shorter periodicities that are less significant [13].

The cross-wavelet analysis further demonstrated the association between climate variables and malaria incidence. The results showed intermittent temporal cycles between temperature and malaria incidence at different window period ranging from annual to 1.5-year cycles in Lagos State as revealed within the 8-16-month band of the cross-wavelet power spectrum (Figure 7a). This association describes the fluctuating interaction between temperature and malaria incidence. Similar temporal association was observed between rainfall and malaria incidence in Lagos State within the 8-16-month band. The dominance of the 8–16-month band suggests that seasonal and inter-annual temperature variability modulated sustained control over malaria incidence rather than short-term fluctuations. In contrast, rainfall exhibited stronger coherence with malaria incidence within the same band, highlighting its continued relevance for malaria risk even in low-transmission settings.

The result of the wavelet transform coherence between the temperature and malaria incidence showed the absence of strong coherence at shorter timescales indicating that sub-seasonal temperature variability alone is insufficient to drive detectable changes in malaria incidence, likely due to offsetting by other ecological and socio-environmental factors such as rainfall patterns, vector control interventions, and healthcare-seeking behavior in Lagos State. Our analysis showed that the rainfall patterns demonstrated a strong coherence with malaria incidence as demonstrated by the arrows in Figure 5 (b), suggesting that it modulated both short-term and long-term influence on malaria transmission in Lagos State. Conversely, weak coherence at longer timescales between the malaria transmission and temperature may reflect the influence of non-climatic drivers that may include intervention scale-up, and other factors not considered in this study, which can obscure long-term climate signals in Lagos State. Results of cross-coherence spectrum indicated that malaria occurrence and temperature were out of phase, whereas they were in phase with rainfall over the three studied climatic zones in Burkina Faso [34]. In Zamfara, the cross-wavelet and coherence analyses reflect a consistent and stable significant association between the climate variables and malaria incidence during the study period. Our findings showed that malaria incidence in Zamfara strongly aligns with temperature and rainfall at the annual timescale. These findings underscore the influence of both rainfall and temperature on malaria transmission in high transmission settings. The alignment of these climatic drivers at similar timescales highlights their joint role in shaping malaria seasonality and inter-annual variability and supports the integration of climate-informed indicators into malaria surveillance and preparedness strategies.

The consistent and stable association between the temporal cycles of the climate variables and malaria incidence indicate sustained eco-suitable conditions for malaria vector, and continous transmission throughout the year in Zamfara state. This highlights the need for climate-sensitive malaria control strategies in Zamfara, where sustained eco-suitable conditions support year-round transmission. Variability in rainfall and temperature was coherently associated with malaria incidence in an endemic rural dry zone locality, using wavelet approach [13]. Similar results from the study conducted in Kano and Jos states of Nigeria highlighted strong association between malaria and climatic factors in high-burden context [35]. The analysis emphasized robust coherence structures, supporting the role of climate variability in shaping transmission dynamics in high-burden contexts.

The phase analysis revealed distinct lead–lag relationships across the two states. Rainfall and temperature patterns in Lagos State demonstrated distinct relationship with malaria incidence. The fluctuating and inconsistent lag between malaria incidence and temperature in Lagos illustrates the weak dominance of temperature on malaria transmission in low-endemic settings. In contrast, the rainfall pattern demonstrated a consistent phase alignment with malaria transmission and an intermittent lead-lag relationship where it consistently leads malaria by approximately one month signifying the influence of rainfall on malaria transmission in Lagos State. These findings mirror results from Sri Lanka [13], and the Brazilian Amazon [19], where rainfall-driven coherence with malaria transmission has been repeatedly observed. Conversely, the fluctuating lead–lag relationship between temperature and malaria incidence in Lagos State is consistent with the less stable temperature effects reported in these regions. These findings suggests that continuous effort to reduce the gains of rainfall to the mosquito population should be geared up to reduce the mosquito breeding sites alongside other control interventions to reach elimination stage in Lagos State.

In Zamfara, the phase analysis revealed that the monthly malaria incidence lagged the temperature by approximately four months on average. This lag suggests why the monthly malaria incidence and temperature in Zamfara state are out of phase as shown in Figure 8(a). This lag is consistent with the observed lags from other studies between temperature and malaria incidence [35–37], which possibly was responsible for the weak correlation between malaria and temperature within the state. The lag between the temperature and malaria incidence is unlikely to reveal biological delays caused by the development of some of the biological traits of the female Anopheles mosquitoes, or the *Plasmodium* parasite. It is presumably a reflection of unsuitable environmental conditions during the hottest months in the year as reflected by peaks in the phase difference.

In particular, temperature peaks usually occur between March and May in Zamfara (for more information, see S1 File). This peak period corresponds to the late dry season in Zamfara state when the monthly mean temperature reaches as high as 35^0^C. The temperature values at these periods exceed the optimal temperature range condition for mosquito survival, breeding site availability and intense malaria transmission [38]. Consequently, malaria incidence does not reach its peak during these months. The observed lag may therefore be attributed to ecological constraints imposed by extreme heats, rather than to delays in biological processes associated with the female Anopheles mosquitoes or the malaria parasites.

High temperature shortens the lifespan of adult mosquitoes, reduces the survival probability of immature mosquitoes, and also leads to dryness of breeding sites thus limiting the availability of space to lay eggs which altogether reduce vector abundance [39, 40]. Nevertheless, as the temperature tends towards the optimal range with the resumption of rainfall, breeding sites are replenished and mosquito survival improves. The temperature values corresponding to when malaria peaks are within the optimal range for malaria transmission as shown in Table S1B. For more information, see S1 File. Consequently, the mean 4-month lag reflects the ecological rebound of mosquito population following the hottest period rather than developmental or incubation delays. This interpretation highlights the importance of ecological context when interpreting climate–malaria lag relationships.

Phase analysis underscores the importance of tailoring malaria control strategies to local epidemiological contexts. In low-burden states, sustaining interventions that weaken climate forcing is critical, while in high-burden states, climate-adaptive surveillance and targeted interventions are needed to anticipate rainfall-driven peaks. At the national level, these regional insights reinforce the importance of integrating climate variability into Nigeria’s malaria control framework. By recognizing that rainfall and temperature exert distinct influences across ecological zones, policymakers can design more flexible, climate-adaptive strategies that align with local transmission patterns. Such an approach would strengthen early warning systems, optimize resource allocation, and enhance the effectiveness of interventions, ultimately advancing Nigeria’s progress toward malaria elimination.

Programatically, our findings outline evidence-based framework to guide climate-adaptive intervention timing. In Zamfara state, extreme heat between March and May as shown in the temperature profile (for more information, see S1 File) may reduce use of LLINs as also reported in [41], indicating that mass distribution before and during these periods, within same year, may be less effective. The start of rain comes with a cooling effect which may facilitates good weather condition that encourages LLIN utilization. Correspondingly, LLIN distribution campaigns conducted in June or July, prior to peak rainfall and peak malaria incidence typically observed between August and October, may enhance intervention effectiveness. Coupled with other climate-sensitive control interventions (for example, seasonal malaria chemo-prevention), such campaigns should be repeated at intervals of no more than three years, in alignment with the observed multi-annual cycles of malaria incidence, to effectively mask malaria risk in Zamafara state. This implementation strategy could be employed in other high transmission states of Nigeria to mitigate malaria risk.

Altogether, these findings emphasise the complex and multi-scale influence of rainfall on malaria transmission in Zamfara State. They highlight the need for climate-informed surveillance systems and adaptive intervention strategies that anticipate both short-term and long-term rainfall-driven transmission cycles. When considered alongside the Lagos analysis, the results reveal both regional similarities—such as the dominant annual cycles—and important differences, including the lagged response in Lagos versus the emergence of semi-annual and biennial cycles in Zamfara. This comparative perspective underscores the necessity of tailoring malaria control strategies to local climatic contexts, ensuring that interventions remain responsive to region-specific transmission dynamics.

## Limitations of the study

This study did not explicitly account the effects of control interventions, thus limiting ability to quantify how these interventions distort the observed temporal cycles in the study areas. Also, variations in intervention coverage, surveillance, and demographic dynamics were not explicitly modeled, which constrain causal interpretation of climate-malaria response. We only used the monthly malaria incidence data not accounting for how climatic factors influence prevalence of malaria across the states.

Additionally, the wavelet-based techniques employed in this study identified localized association rather than mechanistic linkage, limiting the ability of the study to integrate malaria transmission dynamics and its interaction with climatic factors. Future work that integrates mechanistic transmission models with wavelet-derived scale information would strengthen inference and policy relevance of informing early-warning systems.

## Conclusion

The wavelet analysis of the time-dependent relationship between each climate variable and malaria incidence reveals a lead-lag relationship which provides information on intervention timing to support evidence-based decision-making towards control and elimination of malaria across various burden zones. The findings in this study essentially advance our understanding of the temporal characteristics of rainfall and temperature, and their association and relationship with malaria incidence in low- and high-burden states of Nigeria, offering profound insights to improve predictive endemic models that can quantitatively inform sub-national malaria strategies and decisions concomitant to different transmission settings.

Overall, the contrasting malaria–climate relationships observed in Lagos and Zamfara States underscore the heterogeneity of malaria transmission dynamics across Nigeria. In low-burden urban settings, malaria transmission appears increasingly decoupled from climatic variability, while in high-burden rural settings, climate remains a dominant driver of transmission. These findings support the need for context-specific malaria control strategies that integrate climatic information with local epidemiological and environmental conditions to improve surveillance, intervention timing, and progress toward control and elimination of malaria transmission.

## Data Availability

The dataset used for this study is not readily available to the public however, access can be given upon reasonable request from the NMEP through https://nmdrnigeria.ng/dhis-web-common/ security/login.action

https://nmdrnigeria.ng/dhis-web-common/security/login.action

## Supporting information

**S1 File. Cross-epidemiological assessment of seasonal peaks (2015–2024). Table S1A**. Annual peak months of malaria incidence, temperature, and rainfall in Lagos state (2015–2024). **Table S1B**. Annual peak months of malaria incidence, temperature, and rainfall in Zamfara state (2015–2024). **Fig S1A**. Monthly distribution of (a) temperature (°C) and (b) rainfall (mm) in Lagos state, 2015–2024. **Fig S1B**. Monthly distribution of (a) temperature (°C) and (b) rainfall (mm) in Zamfara state, 2015–2024.

## Acknowledgments

The authors express their sincere appreciation to Mrs. Adeyemi Temitope, the ICAMMDA Administrator, for the assistance rendered in printing some materials required in the course of carrying out this study. Our special appreciation goes to National Malaria Elimination Programme (NMEP) for the provision of access to the data used to carry out this study.

## Notes

### Competing Interest Statement

The authors have declared no competing interest.

### Funding Statement

Yes

### Author Declarations

The datasets used in our study were aggregated/summary data, and any individual-level data had been de-identified prior to use in this study.

## References

1. Phillips MA, Burrows JN, Manyando C, van Huijsduijnen RH, Van Voorhis WC, Wells TNC. Malaria. Nature reviews Disease primers. 2017;3:17050. Available from: https://www.ncbi.nlm.nih.gov/pubmed/28770814. doi:10.1038/nrdp.2017.50.

2. National Malaria Elimination Programme, Federal Ministry of Health, Nigeria. National Malaria Strategic Plan 2021–2025; 2021. Federal Republic of Nigeria National Malaria Elimination Programme strategic plan. Strategic plan document. Available from: https://extranet.who.int/cpcd/sites/default/files/public_file_repository/NGA_Nigeria_National-Strategic-Plan-Malaria_2021-2025.pdf.

3. Caminade C, Ayala D, de Chevigny T, Ngou O, Tchouatieu A, Girond F, et al. Climate change and malaria control: a call to urgent action from Africa’s frontlines. Malaria Journal. 2025 06;24. doi:10.1186/s12936-025-05431-5.

4. World Health Organization. World malaria report 2024: addressing inequity in the global malaria response. Geneva: World Health Organization; 2024. Available from: https://www.who.int/publications/i/item/9789240104440.

5. World Health Organization Regional Office for Africa. Report on malaria in Nigeria 2022. Brazzaville: World Health Organization. Regional Office for Africa; 2023. Available from: https://iris.who.int/handle/10665/372274.

6. Nissan H, Ukawuba I, Thomson M. Climate-proofing a malaria eradication strategy. Malaria Journal. 2021 04;20. doi:10.1186/s12936-021-03718-x.

7. Onyango EA, Sahin O, Awiti A, Chu C, Mackey B. An integrated risk and vulnerability assessment framework for climate change and malaria transmission in East Africa. Malaria Journal. 2016 11;15. doi:10.1186/s12936-016-1600-3.

8. Ryan SJ, Lippi CA, Zermoglio F. Shifting transmission risk for malaria in Africa with climate change: a framework for planning and intervention. Malaria Journal. 2020 05;19. Available from: https://malariajournal.biomedcentral.com/articles/10.1186/s12936-020-03224-6. doi:10.1186/s12936-020-03224-6.

9. Lubinda J, Haque U, Bi Y, Hamainza B, Moore AJ. Near-term climate change impacts on sub-national malaria transmission. Scientific Reports. 2021 01;11:751. Available from: https://www.nature.com/articles/s41598-020-80432-9. doi:10.1038/s41598-020-80432-9.

10. Filho WL, May J, May M, Nagy GJ. Climate change and malaria: some recent trends of malaria incidence rates and average annual temperature in selected sub-Saharan African countries from 2000 to 2018. Malaria Journal. 2023 08;22. doi:10.1186/s12936-023-04682-4.

11. Jalloh SW, Malenje B, Imboga H, Hodges MH. Forecasting malaria cases using climate variability in Sierra Leone. Malaria Journal. 2025 05;24. doi:10.1186/s12936-025-05389-4.

12. Singh MP, Rajvanshi H, Bharti PK, Anvikar AR, Lal AA. Time series analysis of malaria cases to assess the impact of various interventions over the last three decades and forecasting malaria in India towards the 2030 elimination goals. Malaria Journal. 2024 02;23. doi:10.1186/s12936-024-04872-8.

13. Mahendran R, Pathirana S, Piyatilake ITS, Perera SSN, Weerasinghe MC. Assessment of environmental variability on malaria transmission in a malaria-endemic rural dry zone locality of Sri Lanka: The wavelet approach. PLOS ONE. 2020 02;15:e0228540. doi:10.1371/journal.pone.0228540.

14. Torrence C, Compo GP. A Practical Guide to Wavelet Analysis. Bulletin of the American Meteorological Society. 1998;79(1):61–78. Available from: https://journals.ametsoc.org/view/journals/bams/79/1/1520-0477_1998_079_0061_apgtwa_2_0_co_2.xml. doi:10.1175/1520-0477(1998)079¡0061:APGTWA¿2.0.CO;2.

15. Cazelles B, Chavez M, Magny GCd, Guégan JF, Hales S. Time-dependent spectral analysis of epidemiological time-series with wavelets. Journal of The Royal Society Interface. 2007 02;4:625–36. doi:10.1098/rsif.2007.0212.

16. Rhif M, Ben Abbes A, Farah I, Martínez B, Sang Y. Wavelet Transform Application for/in Non-Stationary Time-Series Analysis: A Review. Applied Sciences. 2019 03;9:1345. doi:10.3390/app9071345.

17. Kyaw MH, Spinardi JR, Jagun O, Villalobos CF, Kapetanakis V, Sharf-Williams R, et al. Descriptive analysis to assess seasonal patterns of COVID-19 and influenza in low-income and middle-income countries in Asia, the Middle East and Latin America. BMJ Open. 2024 01;14:e081019–9. Available from: https://www.ncbi.nlm.nih.gov/pmc/articles/PMC10831443/#:~:text=Two%20countries%20exhibited%20a%20similar. doi:10.1136/bmjopen-2023-081019.

18. Martin-Makowka A, Nyawanda BO, Beloconi A, Bigogo G, Khagayi S, Munga S, et al. Variation of malaria dynamics and its relationship to climate in western Kenya during 2008–2019: a wavelet approach. eLife. 2025. doi:10.7554/elife.103597.1.

19. Valiati NCM, Rice B, D. Disentangling the seasonality effects of malaria transmission in the Brazilian Amazon basin. Royal Society Open Science. 2024 07;11. doi:10.1098/rsos.231764.

20. Oniyelu DO, Folorunsho O, Adewole L, Bakare EA, Okoronkwo C, Eze N. Time series analysis of malaria in pregnancy, using wavelet and SARIMAX models. PLoS ONE. 2025 08;20:e0328888–8. doi:10.1371/journal.pone.0328888.

21. Bakare EA, Mogbojuri OA, Oniyelu DO, Abidemi A, Daniel DO, Olasupo II, et al. Time series modelling and forecasting of mpox incidence and mortality in Nigeria. BMC Infectious Diseases. 2025 06;25. doi:10.1186/s12879-025-11174-0.

22. Omogunloye OG, Abiodun OE, Olunlade OA, Epuh EE, Odumosu JO, Asikolo I. Modeling Malaria Prevalence Rate in Lagos State Using Multivariate Environmental Variations. Geoinformatics FCE CTU. 2018 08;17:61. doi:10.14311/gi.17.1.5.

23. Abdullahi K, Shinkafi SA, Aguh BI, Bilbis LS, Danladi A, Mohammad BA, et al. Malaria knowledge, attitude, perception and practices in Gusau Metropolis, Zamfara State (a preliminary report II). International Journal of Science for Global Sustainability. 2021;6:8–8. doi:10.57233/ijsgs.v6i4.57.

24. Maigemu AY. The Role of Poverty and Household Economic Conditions to the Treatment of Malaria in Zamfara State North West Nigeria. Journal of Social and Development Sciences. 2015 06;6:30–6. doi:10.22610/jsds.v6i2.839.

25. Statista Research Department. Number of people living in Lagos, Nigeria; 2025. Accessed: 2025-12-01. https://www.statista.com/statistics/1308467/population-of-lagos-nigeria/.

26. Adegbenga Adeonipekun P, Adeyemi Adeniyi T, Mateawo J, Agbalaya B. The Late Quaternary vegetational and environmental history of western tropical Africa: the eastern Benin Basin, Lagos, Nigeria. Geology, Geophysics and Environment. 2017;43:277. doi:10.7494/geol.2017.43.4.277.

27. Lagos State Government. Health Services; 2025. Accessed: 2025-12-01. https://lagosstate.gov.ng/news/Health%20Services/view/67c73079984d6e2dadbf8c95.

28. Usman R, Umar AA, Gidado S, Gobir AA, Obi IF, Ajayi I, et al. Predictors of malaria Rapid Diagnostic Tests’ utilisation among healthcare workers in Zamfara State. PLOS ONE. 2018 12;13:e0200856. doi:10.1371/journal.pone.0200856.

29. World Weather Online. World Weather Online; 2024. Accessed: 2024-XX-XX. https://www.worldweatheronline.com/.

30. Anupong S, Modchang C, Chadsuthi S. Seasonal patterns of influenza incidence and the influence of meteorological and air pollution factors in Thailand during 2009–2019. Heliyon. 2024 09;10:e36703. doi:10.1016/j.heliyon.2024.e36703.

31. Omata K, Shimazaki A. Wavelet analysis of COVID-19 pandemic. Journal of Advanced Simulation in Science and Engineering. 2023 01;10:214–20. Available from: https://www.jstage.jst.go.jp/article/jasse/10/2/10_214/_article. doi:10.15748/jasse.10.214.

32. Roesch A, Schmidbauer H. WaveletComp: Computational Wavelet Analysis; 2018. R package version 1.1. Available from: https://CRAN.R-project.org/package=WaveletComp.

33. Grillet ME, Souki ME, Laguna F, León JB. The periodicity of Plasmodium vivax and Plasmodium falciparum in Venezuela. Acta Tropica. 2014 01;129:52–60. doi:10.1016/j.actatropica.2013.10.007.

34. Traoré N, Millogo O, Sié A, Vounatsou P. Impact of Climate Variability and Interventions on Malaria Incidence and Forecasting in Burkina Faso. International Journal of Environmental Research and Public Health. 2024 11;21:1487–7. Available from: https://pmc.ncbi.nlm.nih.gov/articles/PMC11593955/.. doi:10.3390/ijerph21111487.

35. Akinbobola A, Hamisu S. Malaria and Climate Variability in Two Northern Stations of Nigeria. American Journal of Climate Change. 2022;11:59–78. doi:10.4236/ajcc.2022.112004.

36. Egbom SE, Nduka FO, Nzeako SO, Nwafor GO, Bartholomew DC, Nwaigwe CC, et al. Climatic and governance determinants of malaria transmission in Rivers State, Nigeria. Scientific Reports. 2026 01. doi:10.1038/s41598-026-35029-z.

37. Sena L, Deressa W, Ali A. Correlation of climate variability and malaria: A retrospective comparative study, Southwest Ethiopia. Ethiopian Journal of Health Sciences. 2015 04;25:129–9. doi:10.4314/ejhs.v25i2.5.

38. Mordecai EA, Paaijmans KP, Johnson LR, Balzer C, Ben-Horin T, de Moor E, et al. Optimal temperature for malaria transmission is dramatically lower than previously predicted. Ecology Letters. 2012 10;16:22–30. doi:10.1111/ele.12015.

39. Athni TS, Childs ML, Glidden CK, Mordecai EA. Temperature dependence of mosquitoes: Comparing mechanistic and machine learning approaches. PLoS neglected tropical diseases. 2024 09;18:e0012488–8. doi:10.1371/journal.pntd.0012488.

40. Barr JS, Martin LE, Tate AT, Hillyer JF. Warmer environmental temperature accelerates aging in mosquitoes, decreasing longevity and worsening infection outcomes. Immunity and Ageing. 2024 09;21. doi:10.1186/s12979-024-00465-w.

41. National Malaria Elimination Programme (NMEP), National Population Commission (NPC), ICF. Nigeria Malaria Indicator Survey 2021 Final Report; 2022. Abuja, Nigeria and Rockville, MD, USA. Government report.

